# Early detection of acute kidney injury in critically ill children: Predictive value of renal arterial Doppler assessment

**DOI:** 10.1101/2022.04.20.22274078

**Authors:** Aline Vaconcelos de Carvalho, Isabel de Siqueira Ferraz, Fabiane Mendes de Souza, Marcelo Barciela Brandão, Roberto José Negrão Nogueira, Daniela Fernanda Santos Alves, Tiago H. de Souza

**Affiliations:** Pediatric Intensive Care Unit, Department of Pediatrics, Clinics Hospital of the State University of Campinas (UNICAMP), Campinas, SP, Brazil; Department of Pediatrics, School of Medicine São Leopoldo Mandic, Campinas, SP, Brazil; School of Nursing, State University of Campinas (UNICAMP), Campinas, SP, Brazil

**Keywords:** acute kidney injury, renal failure, doppler ultrasonography, renal resistive index, renal pulsatility index, children, pediatric intensive care unit

## Abstract

**Objective:** Renal resistive index (RRI) and renal pulsatility index (RPI) are Doppler-based variables proposed to assess renal perfusion at the bedside in critically ill patients. This study aimed to assess the accuracy of such variables to predict acute kidney injury (AKI) in mechanically ventilated children.

**Design:** Prospective single-center observational study

**Setting:** Pediatric intensive care unit of a quaternary care teaching hospital.

**Patients:** 84 children under controlled ventilation (median age of 5.1 months and weight of 6.6 kg).

**Interventions:** Consecutive children underwent renal Doppler ultrasound examination within 24 hours of invasive mechanical ventilation. Renal resistive index (RRI) and renal pulsatility index (RPI) were measured. The primary outcome was severe AKI (KDIGO stage 2 or 3) on day 3. Secondary outcomes included the persistence of severe AKI on day 5.

**Results:** On day 3, 22 patients were classified as having AKI (any stage), of which 12 were severe. RRI could effectively predict severe AKI (area under the ROC curve [AUC] 0.94; 95% CI 0.86 – 0.98; *p* < 0.001) as well as RPI (AUC 0.86; 95% CI 0.76 – 0.92; *p* < 0.001). The AUC of the IRR was significantly greater than that obtained from the RPI (*p* = 0.023). The optimal cutoff for RRI was 0.85 (sensitivity, 91.7%; specificity, 84.7%; positive predictive value, 50.0%; and negative predictive value 98.4%). Similar results were obtained when the accuracy to predict AKI on day 5 was assessed. Significant correlations were observed between RRI and estimated glomerular filtration rate at enrollment (ρ = -0.495, *p*<0.001) and on day 3 (ρ = -0.467, *p* <0.001).

**Conclusions:** Renal Doppler ultrasound may be a promising tool to predict AKI in critically ill children under invasive mechanical ventilation.

## INTRODUCTION

Acute kidney injury (AKI) is highly prevalent among hospitalized children, with incidence rates ranging from 5% in non-critically ill children to 50% in those admitted to pediatric intensive care units (PICU) (1). This disease is strongly associated with worse outcomes, including increased mortality, increased use of mechanical ventilation, and prolonged PICU stay (2). Hypoperfusional states (prerenal injury) and intrinsic renal diseases are important causes of AKI in the PICU, the latter being mainly caused by the transformation of prerenal AKI into acute tubular necrosis (ATN) after prolonged hypoperfusion in a pathophysiological continuum (3). With this, early recognition of AKI is essential to promptly initiate supportive care aimed at restoring renal perfusion, which may prevent or attenuate ATN (4). However, accurate and early diagnoses of AKI is one of the greatest challenges faced daily by pediatric intensivists.

Currently, the most widely used criteria for diagnosing and staging AKI is the Kidney Disease: Improving Overall Outcomes (KDIGO) criteria, which is based on acute changes in serum creatinine (sCr) and/or reduced urine output (UO) (5). These criteria have limitations, especially in children. Significant sCr elevations can be delayed 24 to 48 hours after a renal insult and are commonly seen only after at least 50% of renal function loss. In addition, children may have very low sCr levels at baseline, which can make it difficult to identify relative increases when values remain within normal ranges. To overcome these limitations, point-of-care ultrasonography has been proposed as a reliable and safe method to both diagnose and predict the occurrence of AKI in ICU patients.

Renal arterial Doppler-based parameters, such as renal resistive index (RRI) or renal pulsatility index (RPI), are rapid, noninvasive, and repeatable variables that may be promising for early AKI detection. Because it reflects resistance to blood flow, lower RRI values are associated with better renal perfusion, while increased values are associated with progression to ATN (6, 7). However, there is a lack of evidence supporting the use of this technique in PICU. Therefore, this study aims to assess the accuracy of renal arterial Doppler ultrasound (RDU) to predict AKI in mechanically ventilated children. We also evaluated the correlation between the variables obtained by Doppler measurements (RRI and RPI) and some variables of interest, such as diuretic score, urine output, fluid balance, and cardiac index.

## METHODS

### Study design, subjects, and setting

This prospective cohort study was conducted in the PICU of the Clinics Hospital of the State University of Campinas (UNICAMP), Sao Paulo, Brazil, between May and October 2021. The study was approved by the local institutional review board (UNICAMP’s Research and Ethics Committee, approval #44357421.7.0000.5404), and written informed consent was obtained from the participants’ legal guardians.

All patients during the first 24 hours of invasive mechanical ventilation were consecutively assessed for eligibility. Patients were included if they presented the following criteria: 1) normal blood pressure and heart rate for age (8); 2) more than 1 hour without any change in vasoactive agents rate infusion or infusion of fluids to volume expansion; 3) adequate vascular filling proved by a respiratory variation in pulse pressure or peak velocity of aortic blood flow < 13%; 4) serum pH between 7.35 – 7.45; and 5) sinus rhythm. Exclusion criteria were as follows: 1) skin lesions or bandages at the sites of ultrasound or echocardiography exams; 2) known preexistent chronic kidney dysfunction or renal artery stenosis; 3) expected PICU length of stay < 48h; 4) score of Comfort-B scale ≥ 23; 5) death occurring within 72h after inclusion; and 6) unavailability of ultrasound operators.

### Ultrasound Doppler and echocardiography measurements

Doppler ultrasonography and echocardiography were performed at the patient’s bedside with a 2.7-8 MHz phased array probe and/or a 3.5-10.0 linear array probe (Vivid Q; GE Healthcare, Tirat Carmel, Israel). Since children’s renal arterial blood flow on the arcuate or interlobar arteries are difficult to assess, we decided to perform the Doppler measurements on the segmental arteries of the right kidney. The choice of transducer and frequency adjustment was made to obtain the best visualization of the segmental arteries on color Doppler ultrasound. The RRI was calculated as follows: (peak systolic velocity – end-diastolic velocity) / peak systolic velocity. While the RPI was calculated as: (peak systolic velocity – end-diastolic velocity) / mean velocity (***Figure 1***). Three measurements were performed and averaged to obtain the mean RRI and RPI values. Cardiac index and respiratory variation in aorta blood flow peak velocity (ΔVpeak) were determined by transthoracic echocardiography as previously described (9).

**Fig.1.**
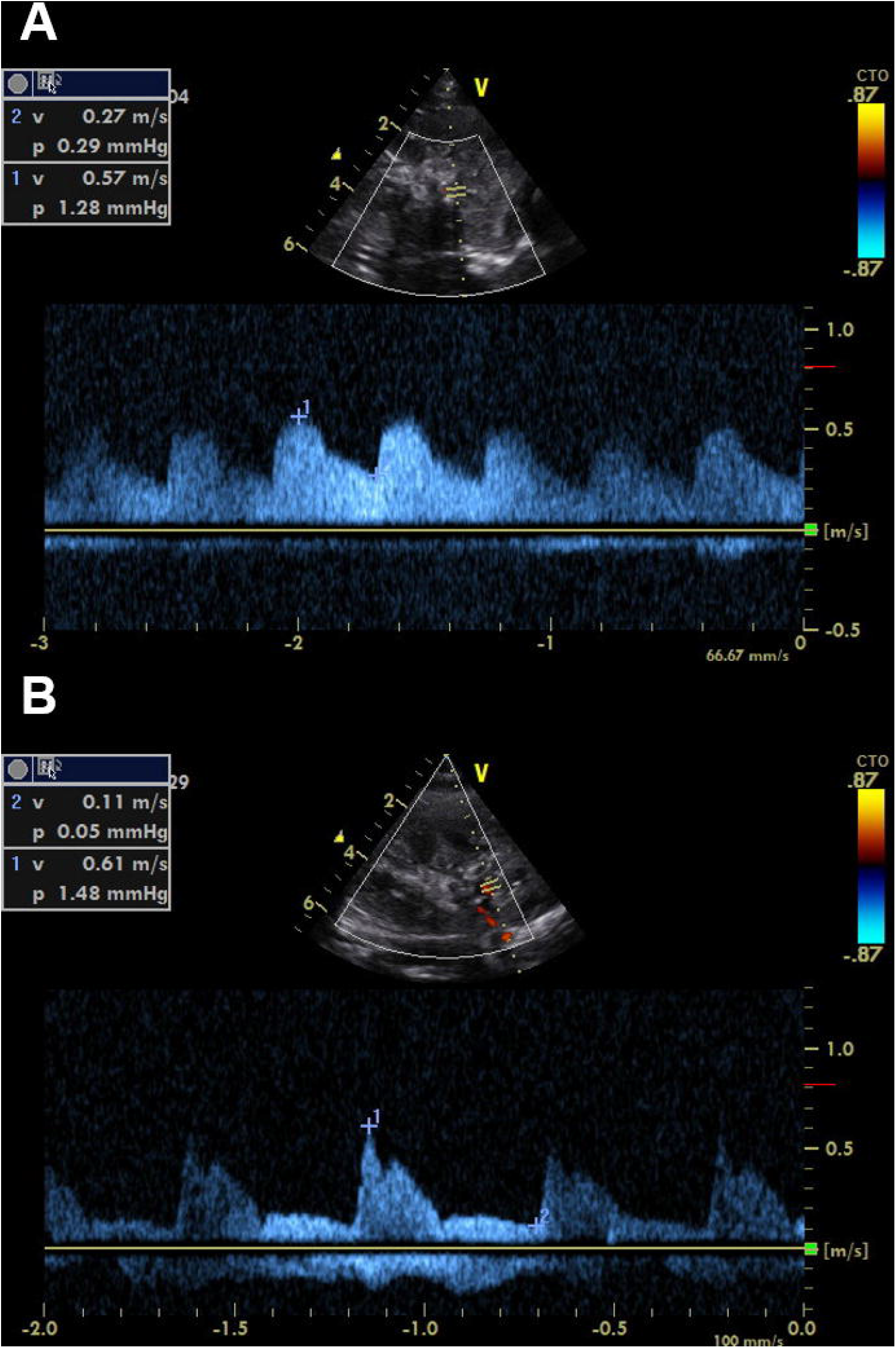
Renal arterial Doppler ultrasonography showing a renal resistance index of 0.58 (A) and 0.82 (B).

Ultrasound and echocardiographic examinations were preferably performed by a qualified pediatric ultrasound instructor of the Brazilian Society of Intensive Care Medicine, with 7 years of experience in pediatric point-of-care ultrasound (POCUS). When the experienced operator was not available, ultrasound examinations were performed by two novice operators who were trained prior to the study. All examinations performed by novice operators were reviewed by the experienced operator.

### Data Collection

At inclusion (D1), the following data were collected:

- Demographic data: age; sex; weight and stature at PICU admission; main diagnosis; and the pediatric index of mortality 2 score.
- Hemodynamic variables: CI; ΔVpeak; heart rate; and blood pressure;
- Ventilatory variables: mean airway pressure; positive end-expiratory pressure (PEEP); peripheral arterial oxygen saturation; and respiratory rate.
- Laboratory variables: SCr; arterial blood gases and serum lactate.
- Doppler-based variables: RRI and RPI.

Then, every morning, urine output, net fluid balance, and diuretic score were recorded until the third day (D3). The diuretic score was calculated using the following formula: furosemide daily dose (mg/kg) plus a tenth of chlorothiazide daily dose (mg/kg). Patients who had a diagnosis of AKI (any stage) on D3 had the aforementioned variables collected until the fifth day (D5).

### Definitions

The occurrence of AKI and its severity was defined according to the KDIGO criteria (8). When available, the baseline sCr was considered as the lowest value in the 3 months prior to PICU admission, otherwise, it was calculated using the Schwartz equation (10).

### Outcomes

The primary outcome was the occurrence of severe AKI on D3, defined as stage 2 or 3 of the KDIGO criteria. Secondary outcomes include: 1) persistent AKI on D5; 2) occurrence of AKI stage 1; 3) renal replacement therapy; 4) diuretic score, urine output; and fluid overload (calculated as the net difference between all inputs and outputs divided by admission weight, expressed as a percentage) on D3 and D5.

### Statistical Analysis

Statistical analysis was performed using MedCalc Statistical Software version 19.8 (MedCalc Software Ltd, Ostend, Belgium). The Kolmogorov-Smirnov and Shapiro-Wilk tests were used to assess the normality of the data distribution. Continuous variables were non-normally distributed and were thus described as medians and interquartile range (IQR). Categorical variables were expressed as absolute numbers (%) and were analyzed using either chi-square or Fisher exact test. Continuous variables were compared using Wilcoxon signed-rank test. Intra-rater reliability for RRI and RPI was assessed by calculating the intraclass correlation coefficient (ICC) by using a model for 2-way random single measures (consistency). Receiver Operating Characteristic (ROC) curves were constructed to evaluate the diagnostic value of RRI and RPI for predicting severe AKI. The optimal cut-off points were defined according to the optimal Youden’s J statistic using univariable analysis. Multivariate logistic regression was fitted to the primary outcome (dependent variable) using a stepwise selection procedure. Independent predictors were entered into the model on the basis of the bivariate analysis (*p* < 0.05) and correlation coefficients between variables (collinearity) lower than 0.5. The Spearman’s correlation coefficient (ρ) was used to estimate and test the link between the variables obtained by Doppler measurements (RRI and RPI) and the following variables of interest: cardiac index, diuretic score, urine output, and fluid overload. P-values <0.05 were considered significant.

The required sample size was calculated to evaluate the primary outcome. We determined a power of 0.80, an alpha of 0.05, and we estimated a prevalence of 15% of severe AKI on D3. To demonstrate that the areas under the ROC curve (AUC) of 0.75 for Doppler-based variables are significantly different from the null hypotheses value 0.5 (meaning no discriminating power) 80 participants were required.

## RESULTS

### Participants

Among the 302 patients admitted to the PICU during the study period, 100 met the inclusion criteria. Sixteen patients were excluded due to a variety of reasons (***Figure 2***), resulting in 84 participants completing the study. Median age and weight at admission were 5.1 months (IQR 1.9 – 24.8) and 6.6 kg (IQR 4.3 – 10.8). Baseline characteristics of participants with and without severe AKI are presented separately in ***Table 1***.

**Fig.2.**
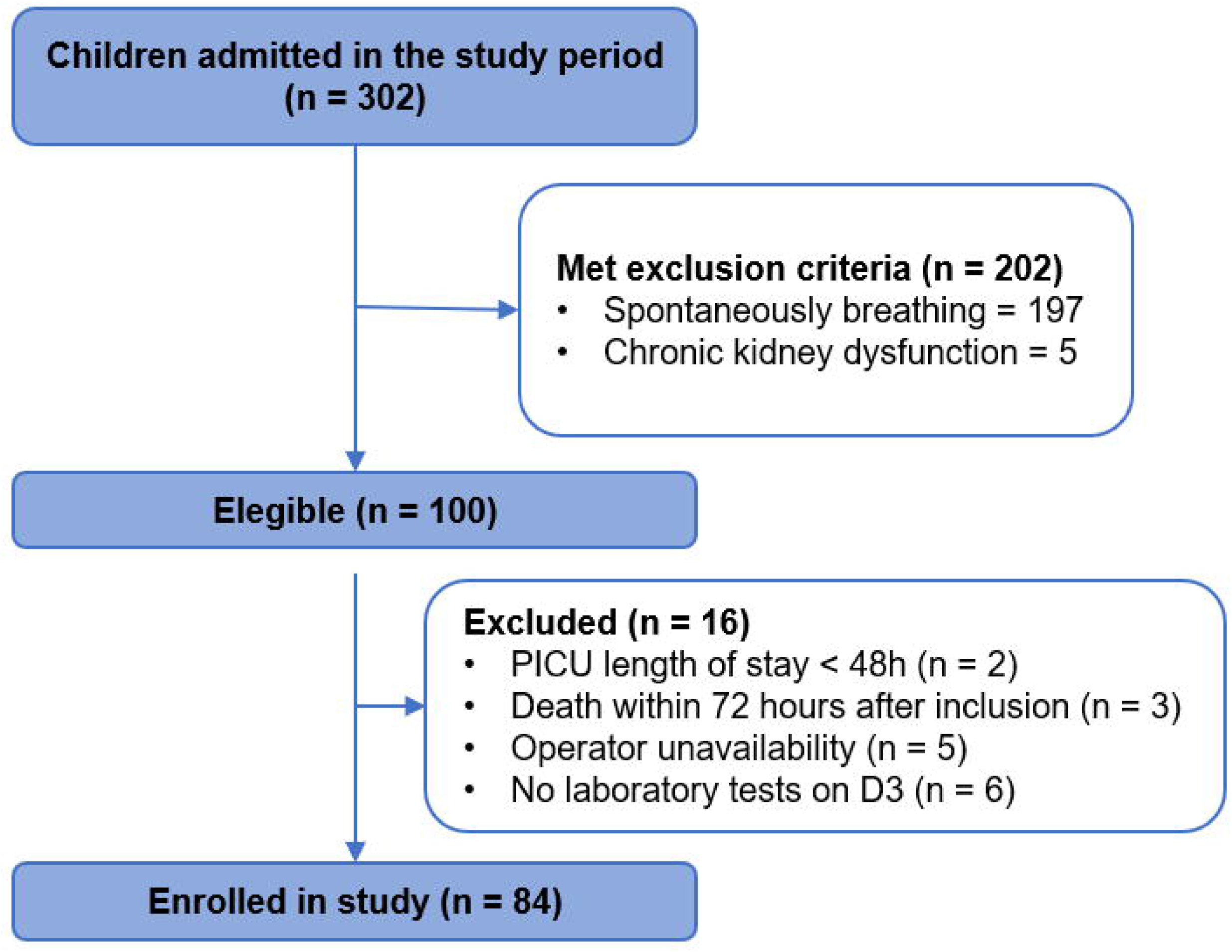
The flow chart of the recruitment process.

**Table 1.**
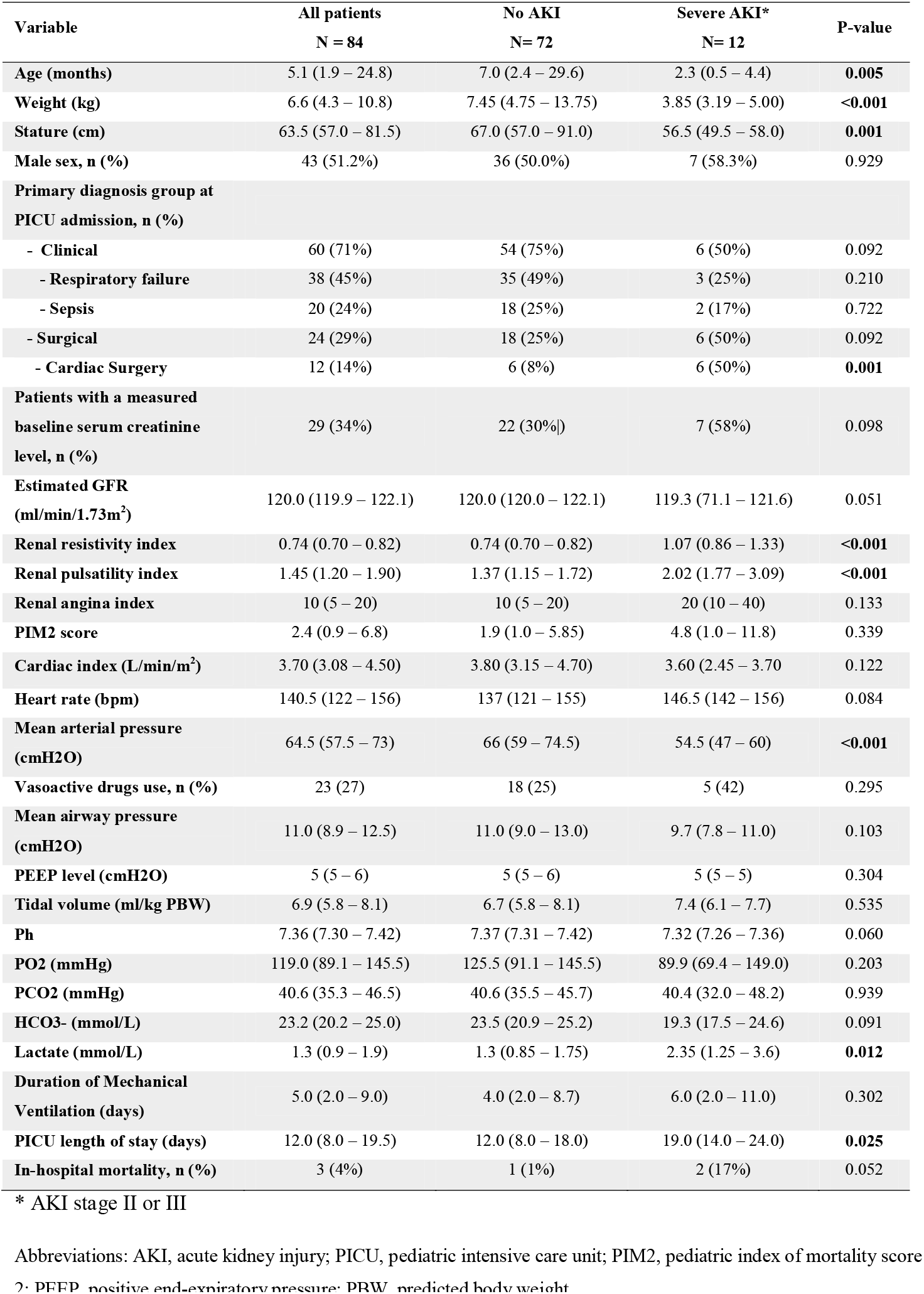
Clinical and demographic characteristics of participants at inclusion according to the presence or absence of severe AKI at D3.

### Acute Kidney Injury

A total of 15 participants had some stage of AKI at inclusion, of which 10 were KDIGO stage 1 and 5 were severe. On D3, 10 subjects were diagnosed with AKI stage 1 and 12 with severe AKI. Of these, 10 participants remained alive on D5, 7 of which had persistent severe AKI. Six participants required peritoneal dialysis due to oliguria or anuria, 5 of whom were in post-operative cardiac surgery care.

### Renal Doppler ultrasound

The vast majority of ultrasound examinations were performed by the experienced operator (76/84). The intra-operator reliability was 0.99 (95% CI 0.98 - 0.99) for RRI and 0.86 (95% CI 0.80 - 0.90) for PI.

Both RRI and RPI were predictors of severe AKI on D3, presenting an AUC of 0.94 (95% CI 0.86 – 0.98; *p* < 0.001) and 0.86 (95% CI 0.76 – 0.92; *p* < 0.001), respectively. The AUC of the IRR was significantly greater than that obtained from the RPI (*p* = 0.023). The optimal cut-off for RRI was 0.85, where the sensitivity was 91.7% (95%CI 61.5 – 99.8%), specificity was 84.7% (95%CI 74.3 – 92.1%), PPV was 50.0% (95%CI 28.2 – 71.8%), and NPV was 98.4% (95%CI 91.3 – 100.0%). Similar results were obtained when the accuracy to predict AKI on D5 was assessed. The AUC values are shown in ***Table 2*** and ***Figure 3***.

**Table 2.** Areas under the ROC curve for assessed predictors of severe AKI on day 3 and day 5. Cut point calculated using Youden’s index.

| Performance | Day 3 (N=84) |  | Day 5 (N=82) * |  |
| --- | --- | --- | --- | --- |
|  | RRI | RPI | RRI | RPI |
| **AUROC** | 0.936 (0.861 – 0.978) | 0.858 (0.764 – 0.924) | 0.893 (0.806 – 0.951) | 0.778 (0.673 – 0.862) |
| **Optimal cut-off** | >0.85 | > 1.57 | >0.85 | > 1.57 |
| **Sensitivity (%)** | 91.7% (61.5 – 99.8) | 91.7% (61.5 – 99.8) | 85.71% (42.1 – 99.6) | 85.71% (42.1 – 99.6) |
| **Specificity (%)** | 84.7% (74.3 – 92.1) | 66.7% (54.6 – 77.3) | 81.33% (70.7 – 89.4) | 64.00% (52.1 – 74.8) |
| **PPV** | 50.0% (36.1 – 63.9) | 31.4% (24.1 – 39.9) | 30.0% (19.7 – 42.9) | 18.2% (12.7 – 25.4) |
| **NPV** | 98.4% (90.3 – 99.8) | 98.0% (87.9 – 99.7) | 98.4% (90.8 – 99.7) | 98.0% (88.6 – 99.7) |
| **PLR** | 6.0 (3.4 – 10.6) | 2.75 (1.9 – 4.0) | 4.59 (2.6 – 8.0) | 2.38 (1.6 – 3.7) |
| **NLR** | 0.098 (0.02 – 0.60) | 0.13 (0.02 – 0.80) | 0.18 (0.03 – 1.1) | 0.22 (0.04 – 1.4) |

Abbreviations: AUROC, area under the receiver operating characteristic curve; PPV, positive predictive value; NPV, negative predictive value; PLR, positive likelihood ratio; NLR, negative likelihood ratio.
\* Two participants died before the fifth day and were excluded from this analysis.

**Fig.3.**
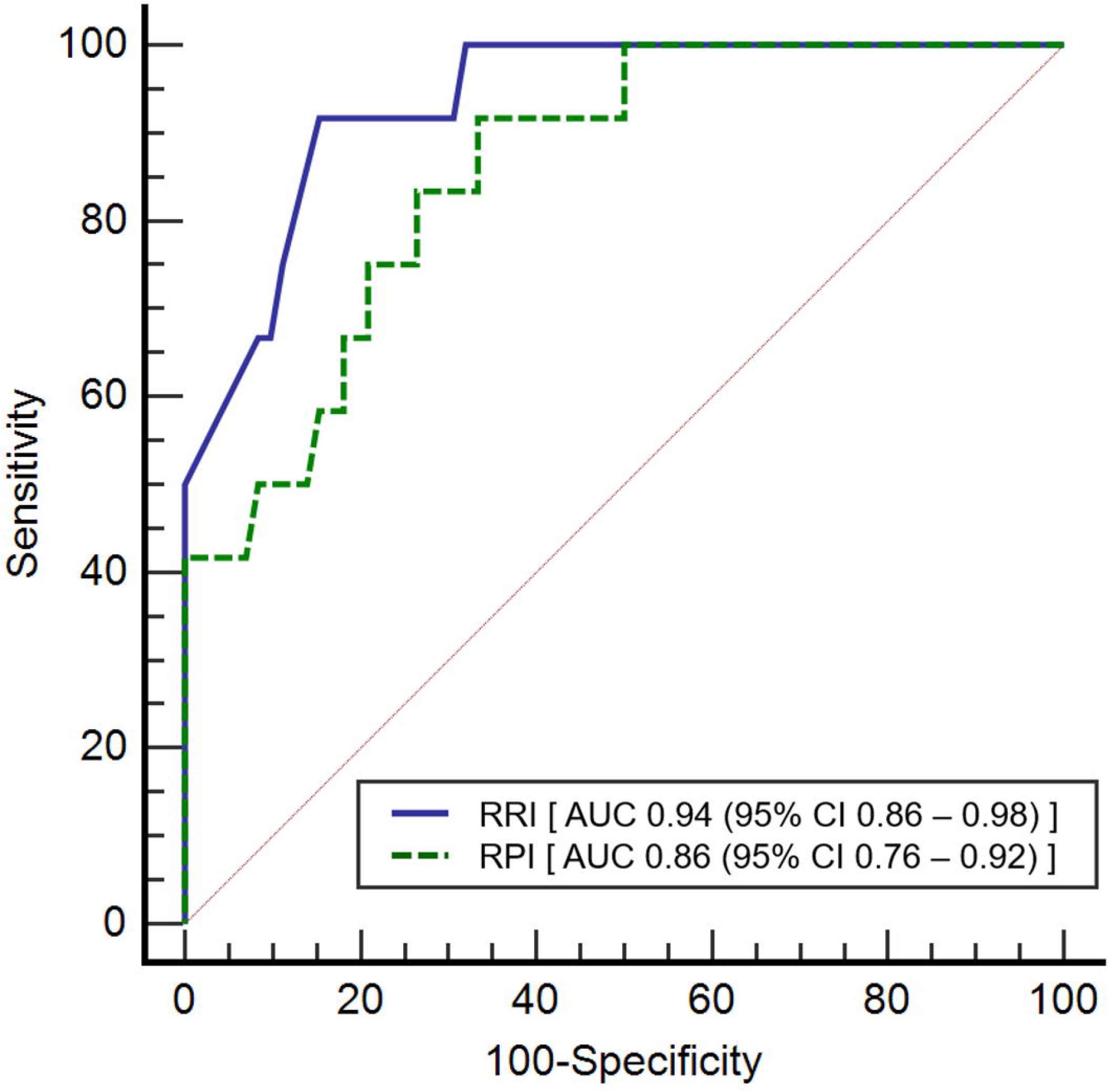
ROC curve for renal resistive index (solid line) and pulsatility index (dashed line) upon inclusion as predictors of severe acute kidney injury on D3.

Univariate analysis showed that AKI on D3 significantly correlated with RRI (ρ = 0.530), RPI (ρ = 0.434), weight (ρ = -0.310), stature (ρ = -0.365), age (ρ = -0.310), urine output on day 1 (ρ = 0.372), estimated GFR (ρ = -0.363), fluid balance on day 1 (ρ = 0.275), and diuretic score on day 1 (ρ = 0.242). No significant correlations were observed between AKI on day 3 and CI or PIM2 score. After assessing for collinearity, the variables age, stature and RPI were excluded from the multivariate logistic regression analysis. After including the other variables, only RRI was retained in the model (OR = 1.13, 95% CI 1.06 – 1.22; *p*<0.001).

Correlation analyzes between the pairs of variables collected at enrollment, D1 and D3, are shown in ***Table 3***. Significant correlations were observed between RRI and estimated glomerular filtration rate at enrollment (ρ = -0.495, *p*<0,001) and on D3 (ρ = -0,467, *p* <0.001). Also, a significant correlation was observed between RRI and diuretic score on D1 (ρ = 0.360, *p*<0.001) and D3 (ρ = 0.388, *p*<0.001). On D1, PI was significantly correlated with estimated glomerular filtration rate (ρ = -0.260, *p*=0.017) and cardiac index (ρ = -0.278, *p*=0.010).

**Table 3.** Correlations between pair of variables evaluated on day 1 and day 3 were tested using Spearman’s correlation coefficient (ρ).

| Pair of variables | Day 1 |  | Day 3 |  |
| --- | --- | --- | --- | --- |
| | $\rho$ | P-value | $\rho$ | P-value |
| IRR and estimated GFR (ml/min/1.73m<sup>2</sup>) | -0,495 * | <**0,001** | -0,467 | <**0,001** |
| IP and estimated GFR (ml/min/1.73m<sup>2</sup>) | -0,260 * | **0,017** | -0,212 | 0,053 |
| IRR and cardiac index (L/min/m<sup>2</sup>) | -0,200 * | 0,067 | - | - |
| IP and cardiac index (L/min/m<sup>2</sup>) | -0,278 * | **0,010** | - | - |
| IRR and urine output (ml/kg/h) | -0,170 | 0,125 | -0,039 | 0,726 |
| IP and urine output (ml/kg/h) | -0,132 | 0,234 | -0,132 | 0,233 |
| IRR and fluid balance (ml/kg) | 0,244 | 0,244 | 0,024 | 0,830 |
| IP and fluid balance (ml/kg) | 0,026 | 0,815 | -0,047 | 0,676 |
| IRR and diuretic score | 0,360 | <**0,001** | 0,388 | <**0,001** |
| IP and diuretic score | 0,164 | 0,139 | 0,184 | 0,096 |

\* Correlation assessed with data collected at enrollment.

## DISCUSSION

The results herein reported revealed that renal Doppler ultrasonography may be a valuable method to predict the occurrence of severe AKI in mechanically ventilated children. During the first day of mechanical ventilation, both RRI and RPI were significantly higher in the group of children who had severe AKI on the third day and high values of AUC were observed (0.936 and 0.858 for RRI and RPI, respectively). It is also noteworthy that higher RRI was significantly correlated with lower urine output and greater use of diuretics. To the best of our knowledge, this is the first study to evaluate the use of renal Doppler-based variables in critically ill children.

However, our results need to be interpreted with caution, as restricted inclusion criteria were applied, which limits its generalization. The decision to include only mechanically ventilated children was made to facilitate the performance of ultrasound exams, since children without sedation might not be collaborative. In addition, this population has an increased risk of developing AKI, which would increase the chance of observing the primary outcome. Thus, the feasibility of the method in spontaneously breathing children still needs to be studied. Also, based on previous studies, we decided to include participants after 1 hour of hemodynamic stabilization, as our objective was to assess the occurrence of AKI on the third day (11, 12). It is possible that renal Doppler ultrasound of hemodynamically unstable patients may be valuable in predicting earlier outcomes or in differentiating prerenal failure from intrinsic renal failure. Further studies are needed to support the use of the method during the restoration of hemodynamic stability in children.

Renal Doppler-based variables represent a rapid and non-invasive method to evaluate kidney hemodynamics. Its main determinants are intrarenal arterial resistance, arterial compliance (i.e., renal interstitial and intra-abdominal pressures), age, and central hemodynamic parameters (13, 14). Early studies evaluated the performance of RRI to distinguish prerenal from ischemic intrinsic azotemia (7, 15, 16). It was observed that adults with parenchymal renal failure usually have RRI > 0.70, while those with pre-renal failure or normal renal function have RRI ranging between 0.50 - 0.71 (13). Differentiating transient from persistent AKI is valuable for treatment decision-making, as prompt restoration of renal perfusion may prevent or attenuate the development of ATN. Intrinsic renal diseases present high intrarenal arterial resistance and/or reduced arterial compliance, which can lead to reduced renal blood flow even when the hemodynamics status has been restored.

Although the role of RRI has been evaluated by several observational studies involving adults, there is no consensus regarding its exact predictive value. One of the first studies on this topic performed in an ICU setting was published in 2006 (11). Similar to our results, Lerolle et al. observed that high RRI obtained in the first hours of care in patients with septic shock were associated with the development of severe renal dysfunction (RIFLE-I or F) on the fifth day. The best cut-off determined was 0.75, with an AUC of 0.85 (95% CI, 0.68 – 0.94), presenting sensibility and specificity of 78% and 77%, respectively (11). In 2015, a meta-analysis of 9 heterogeneous studies, including a total of 176 participants, found that increased RRI or RPI are associated with persistent AKI (17). The pooled sensitivity and specificity were 83% (95%CI, 0.77 – 0.88) and 84% (95% CI, 0.79 – 0.88), respectively, while the summary positive and negative likelihood ratios were 4.9 (95% CI, 2.44 – 9.87) and 0.21 (95% CI, 0.11 – 0.41)(17).

In the present study, the optimal cut-off value found for RRI to predict severe AKI was 0.85, which is higher than the values reported in previous studies involving adults. This was not surprising and two main factors may have contributed to this difference. First, it is well known that RRI in early childhood is considerably higher when compared to older children and the adult population (18, 19). Although the normal range of renal Doppler-based variables has not yet been extensively studied in children, the RRI appears to drop to adult levels only around 4 - 6 years of age (18, 19). This might be explained by developmental renal physiology. Glomerular filtration rate, tubular excretory capacity, and renal blood flow are decreased in neonates and mature during the first years of life (20). In addition, active plasma renin levels are increased and decrease reaching adult levels by 4 - 8 years old (21). Second, we performed measurements of Doppler-based variables at the level of segmental arteries, while other authors evaluated the arcuate or interlobar arteries. This approach was decided as performing an adequate spectral Doppler analysis in smaller arteries would be a challenge. It was observed that values obtained in the segmental arteries are higher than those obtained in interlobar and arcuate arteries (22).

In our study, we did not find a correlation between cardiac index and RRI. The same was observed by Dewitte et al. in adults with septic shock and by Bossard et al. in elderly who underwent cardiac surgery with cardiopulmonary bypass (23, 24). These results suggest that therapies aimed at increasing cardiac output, such as fluid infusion or inotropic agents, may not result in an increase in renal blood flow. In fact, the relationship between macro-hemodynamic parameters and RRI seems to be complex in critical care patients, as intrarenal vasomotility is often altered due to primary disease and pharmacological treatments (25). Some authors have reported a poor inverse correlation between mean arterial pressure and RRI. However, Dewitte et al. observed this correlation only in the group of patients without AKI (23). Therefore, mean arterial pressure and cardiac output cannot be pointed out as the main determinants of RRI in critically ill patients. Moreover, the increased resistance to renal blood flow seen in sustained kidney injury can significantly impair renal perfusion, even when adequate hemodynamic support is provided.

There are some limitations in the present study. First, this is a single-center pilot study including a homogeneous population of critically ill children, which may limit its generalizability. Further studies should evaluate children stratified by age and specific pathologies, such as sepsis, cardiovascular diseases, and post-operative care of major surgeries. Second, almost all ultrasonographic exams were performed by the experienced operator and inter-operator agreement was not evaluated. Since renal Doppler ultrasound is a highly operator-dependent method, determining the minimum theoretical-practical required training to achieve a proper performance is crucial to establish its clinical applicability in children. Third, the ultrasound operator was not blinded to the clinical condition of the participants, although the primary outcome was assessed 3 days later and was based on predefined criteria. Fourth, the criteria used to diagnose AKI have some limitations inherent to the physiological basis of the criteria themselves. However, in the lack of a true gold standard, the KDIGO definition is currently accepted for AKI diagnosis and staging.

## CONCLUSIONS

The present study shows that renal Doppler ultrasonography is feasible and may be a promising tool to predict AKI in critically ill children under invasive mechanical ventilation. Doppler-based variables accurately predict the occurrence of severe AKI and were correlated with urine output and the use of diuretics.

## Data Availability

All data produced in the present study are available upon reasonable request to the authors

## Acknowledgements

Thank you to Carolina Grotta Ramos Telio for her review of the manuscript. We also thank the legal guardians of the participants, attending physicians, pediatric critical care residents, and the nursing staff.

